# Post-Market Surveillance of Six COVID-19 Point-of-Care Tests Using Pre-Omicron and Omicron SARS-CoV-2 Variants

**DOI:** 10.1101/2024.01.05.24300772

**Authors:** Hannah M. Exner, Branden S. J. Gregorchuk, AC-Green Castor, Leandro Crisostomo, Kurt Kolsun, Shayna Giesbrecht, Kerry Dust, David Alexander, Ayooluwa Bolaji, Zoe Quill, Breanne M. Head, Adrienne F. A. Meyers, Paul Sandstrom, Michael G. Becker

**Affiliations:** JC Wilt Infectious Diseases Research Centre, National Microbiology Laboratory Branch, Public Health Agency of Canada, Winnipeg, Manitoba, Canada; Department of Microbiology, University of Manitoba, Winnipeg, Manitoba, Canada; Cadham Provincial Laboratory, Winnipeg, Manitoba, Canada; Office of Population and Public Health, Indigenous Services Canada, Ottawa, Ontario, Canada; Department of Medical Microbiology and Infectious Diseases, University of Manitoba, Winnipeg, Manitoba, Canada

## Abstract

Post-market surveillance of test performance is a critical function of public health agencies and clinical researchers that ensures diagnostics maintain performance characteristics following their regulatory approval. Changes in product quality, manufacturing processes over time, or the evolution of new variants may impact product quality. During the COVID-19 pandemic, a plethora of point-of-care tests (POCTs) were released onto the Canadian market. This study evaluated the performance characteristics of several of the most widely-distributed POCTs in Canada, including four rapid antigen tests (Abbott Panbio, BTNX Rapid Response, SD Biosensor, Quidel QuickVue) and two molecular tests (Abbott ID NOW, Lucira Check IT). All tests were challenged with 149 SARS-CoV-2 clinical positives, including multiple variants up to and including Omicron XBB.1.5, as well as 29 clinical negatives. Results were stratified based on whether the isolate was Omicron or pre-Omicron as well as by RT-qPCR Ct value. The test performance of each POCT was consistent with the manufacturers’ claims and showed no significant decline in clinical performance against any of the variants tested. These findings provide continued confidence in the results of these POCTs as they continue to be used to support decentralized COVID-19 testing. This work demonstrates the essential role of post-market surveillance in ensuring reliability in diagnostic tools.

## INTRODUCTION

During the COVID-19 pandemic, it became clear that an effective public health response required rapid and widespread testing for SARS-CoV-2. Although testing initially relied on laboratory-based reverse transcriptase quantitative PCR (RT-qPCR), decentralized testing quickly became the preferred screening method (1, 2) facilitated through the use of point-of-care tests (POCTs). This shift was accelerated following the emergence of the highly transmissible Omicron variant (Pango lineage B.1.1.529) in December 2021 (3–5) causing unprecedented testing demand that overwhelmed centralized testing facilities (4–6). As of August 2023, 55 POCTs have been authorized for use in Canada through the “*Interim order respecting the importation and sale of medical devices for use in relation to COVID-19*” (7). These devices can be broadly categorized into two groups: molecular tests and rapid antigen detection tests (RADTs). Molecular POCTs detect the presence of SARS-CoV-2-specific nucleic acid sequences through PCR or isothermic amplification techniques such as loop-mediated isothermal amplification (LAMP) (1, 8, 9). Molecular tests generally provide a high level of sensitivity and may detect viral RNA weeks after infection (9–11). In comparison, RADTs detect SARS-CoV-2 surface proteins with decreased sensitivity, particularly when considering asymptomatic cases with lower viral loads (12–14).

Post-market surveillance monitors the performance of POCTs following commercialization and distribution as well as provides independent investigations that assess the manufacturers’ claims regarding test efficacy in real-world settings (15). Ongoing monitoring becomes exceedingly important for POCTs as few quality controls are in place compared to clinical testing performed in centralized test facilities. Test performance may also deviate over time due to changes in manufacturing materials or processes (16, 17), or through viral evolution of test targets. For molecular POCTs, test performance may be impacted by base substitutions at the primer binding sites, which can often be predicted *in silico* (*18*). RADT performance may also be impacted by mutations within target proteins, however, predicting this computationally is more challenging as it is influenced by protein 3D structure. Therefore, post-market surveillance of RADTs relies more heavily on experimentation.

Initial studies evaluating the performance of various RADTs found reduced impaired detection of Omicron variants compared to previously circulating strains (19–21). For example, Osterman et al. reported that multiple RADTs tested had 101-fold lower sensitivity against Omicron BA.1 compared to Delta (21). Although this study may raise concerns surrounding the continued use of these RADTs, multiple subsequent studies demonstrated equivalent sensitivity regardless of variant (22–24). Together, this emphasizes the need to disseminate negative test results in post-market surveillance studies.

Due to the variety of POCTs available globally, tests evaluated in published research often have little to no overlap with the POCTs widely distributed in Canada. Although manufacturers may provide post-market surveillance data regarding performance, these may be based heavily on *in silico* analyses and experimental results may differ (25). Therefore, there is a need for regulatory bodies, clinicians, and public health officials to conduct ongoing aftermarket evaluations and to effectively communicate the results to POCT users. Accordingly, the aim of this post-market surveillance study was to assess the analytical performance of six widely distributed POCTs and to determine if there were differences in the detection of pre-Omicron and Omicron variants.

## MATERIALS AND METHODS

SARS-CoV-2 positive and negative patient samples were used to determine the sensitivity, specificity, and experimental limit of detection (LoD) of each of the COVID-19 POCTs. Clinical samples, reference cycle thresholds (Ct), and lineage data were provided by Cadham Provincial Laboratory (CPL; Manitoba, Canada). Reference Ct values were characterized on remnant universal transport media (UTM) from 149 SARS-CoV-2 positive and 29 negative nasopharyngeal swabs using the Roche 6800 Cobas SARS-CoV-2 test targeting the Open Reading Frame 1a and Envelope genes. Samples were then categorized based on lineage into pre-Omicron (Alpha [B.1.1.7], Delta [B.1.617.2]) or Omicron (BA.1, BA.2, BA.4, BA.5, or XBB.1.5) groups (Table 1). For samples for which lineage data was not available (collected before routine sequencing), lineage was predicted based on the sampling date. No ethics were required under Article 2.4 and Article 2.5 of the *Tri-Council Policy Statement Ethical Conduct for Research Involving Humans* (26) as these were remnant clinical specimens used for assay validation. To minimize sample degradation, each sample was limited to a maximum of two freeze-thaw cycles.

**TABLE 1.**
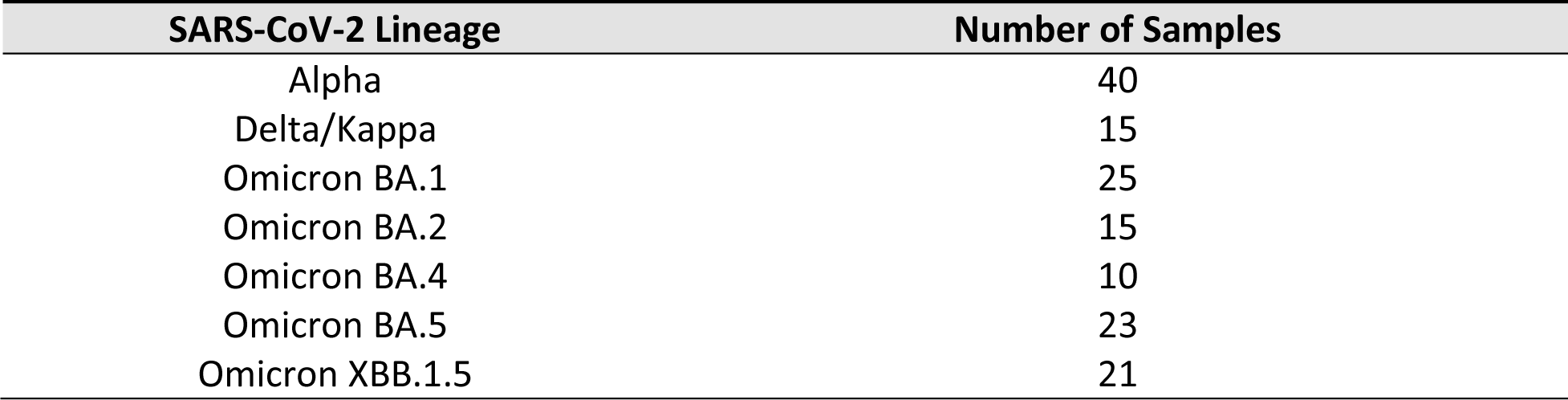
Lineage of SARS-CoV-2 positive clinical samples used in the study.

| SARS-CoV-2 Lineage | Number of Samples |
| --- | --- |
| Alpha | 40 |
| Delta/Kappa | 15 |
| Omicron BA.1 | 25 |
| Omicron BA.2 | 15 |
| Omicron BA.4 | 10 |
| Omicron BA.5 | 23 |
| Omicron XBB.1.5 | 21 |

Six Health Canada-approved COVID-19 POCTs were evaluated in this study; four RADTs including the BTNX Rapid Response® COVID-19 Antigen Rapid Test (BTNX Inc., Canada), the Abbott Panbio™ COVID-19 Ag Rapid Test Device (Abbott Rapid Diagnostics Jena Gmbh, Germany), the SD Biosensor COVID-19 Ag Test (SD Biosensor Inc., South Korea) and the Quidel QuickVue® At-Home OTC COVID-19 Test (Quidel Corporation, United States); as well as two LAMP-based molecular tests: the Abbott ID NOW™ COVID-19 Assay (Abbott Diagnostics Scarborough, Inc., United States) and the Lucira™ Check IT COVID-19 Test Kit (Lucira Health, Inc., United States).

For each POCT, 10 µL of remnant UTM was transferred directly into the assay buffer and subsequently mixed with the manufacturer-provided swab. Each test was then performed according to the manufacturer’s instructions. For RADT results that required visual interpretation, test strips were read independently by three technicians and the consensus was reported. If necessary, invalid tests were repeated until a positive or negative result was encountered.

Data were analyzed using R version 4.2.1 software. Sample Ct values were adjusted to account for the quantity of virus input into the device corresponding to the concentration when eluted into a 3 mL volume. Continuous variables such as Ct are represented as medians and interquartile ranges (IQR). Categorical variables, such as sensitivity and specificity, are reported as percentages with 95% confidence intervals (95% CI) calculated using the Wilson score method. Sensitivity was also determined for each POCT when samples were stratified based on a Ct of ≤ 30 and > 30, i.e., the acceptable LoD outlined by the World Health Organization (WHO) for RADTs (27) and the cut-off typically used in clinical settings and other evaluations (2, 16, 28). For each POCT, the overall and stratified sensitivity values were compared between the pre-Omicron and Omicron groups using a two-tailed Fisher’s Exact Test with an α of 0.05.

Next, based on the guidelines recommended by the Clinical & Laboratory Standards Institute (29), a probit model was constructed for each assay to determine the 95% LoD, representing the estimated concentration at which 95% of positive samples are successfully identified. The 95% CI for each LoD was calculated using the Wald method, then the LoDs were compared between Pre-Omicron and Omicron to determine if the analytical sensitivity of each POCT differed by variant.

## RESULTS

### Performance Characteristics of POCTs with Pre-Omicron and Omicron Variants

A total of 149 SARS-CoV-2 clinical positives with a median Ct value of 30.2 (IQR 26.8–33.8) and 29 SARS-CoV-2 clinical negatives were tested using all six POCTs. Of the positive samples, 94 were determined to be Omicron subvariants (median Ct value 29.7; IQR 27.1–33.5) and the remaining 55 were identified as pre-Omicron (median Ct value 30.0; IQR 26.0–34.4). No false positives were recorded resulting in a specificity of 100% (95% CI, 88–100%) across all tests. Three invalid results were obtained during the study: two with the ID NOW and one with the Check IT.

Overall sensitivity of RADTs ranged from 46-65% compared to 86-98.9% for molecular tests (Table 2). For all tests, sensitivity was similar between Omicron compared to pre-Omicron samples when analyzing all Ct values together. Similarly, when stratifying test results by Ct for all tests, no significant difference in sensitivity was observed between pre-Omicron and Omicron samples with high viral loads (i.e., Ct ≤ 30; Table 2). At low viral loads (i.e., Ct > 30), significant differences in ID NOW (p = 0.021) and QuickVue (p = 0.028) sensitivity was observed between pre-Omicron and Omicron variants.

**Table 2.** Performance characteristics of six COVID-19 point-of-care tests with Pre-Omicron and Omicron samples.

| Test | Ct | Pre-Omicron |  |  | Omicron |  |  | P-value |
| --- | --- | --- | --- | --- | --- | --- | --- | --- |
|  |  | n | Sensitivity (%) 95% CI |  | n | Sensitivity (%) 95% CI |  |  |
| Molecular Tests |  |  |  |  |  |  |  |  |
| ID NOW | Overall | 55 | 86 | (74–92.4) | 94 | 94.7 | (88–97.7) | 0.072 |
|  | ≤30 | 30 | 100 | (89–100) | 41 | 100 | (91.4–100) | 1.000 |
|  | >30 | 25 | 68 | (48–83) | 53 | 90.6 | (80–95.9) | **0.021\*** |
| Check IT | Overall | 55 | 98.2 | (90.4–99.7) | 94 | 98.9 | (94.2–99.8) | 1.000 |
|  | ≤30 | 30 | 100 | (91.4–100) | 41 | 100 | (89–100) | 1.000 |
|  | >30 | 25 | 98.1 | (90.1–99.7) | 53 | 99.3 | (81–99.3) | 0.541 |
| Rapid Antigen Detection Tests |  |  |  |  |  |  |  |  |
| Rapid Response | Overall | 55 | 64 | (50–75) | 94 | 65 | (55–74) | 1.000 |
|  | ≤30 | 30 | 96.7 | (83–99.4) | 41 | 97.6 | (87–99.6) | 1.000 |
|  | >30 | 25 | 24 | (12–43) | 53 | 40 | (28–53) | 0.210 |
| Panbio | Overall | 55 | 51 | (38–64) | 94 | 49 | (39–59) | 0.866 |
|  | ≤30 | 30 | 90.2 | (78–96.1) | 41 | 83 | (66–92.7) | 0.479 |
|  | >30 | 25 | 17 | (9.2–29) | 53 | 12 | (4.2–30) | 0.742 |
| QuickVue | Overall | 55 | 49 | (36–62) | 94 | 55 | (45–65) | 0.499 |
|  | ≤30 | 30 | 87 | (70–94.7) | 41 | 92.7 | (81–97.5) | 0.446 |
|  | >30 | 25 | 4.0 | (0.7–20) | 53 | 26 | (16–40) | **0.028\*** |
| Biosensor | Overall | 55 | 60 | (47–72) | 94 | 47 | (37–57) | 0.130 |
|  | ≤30 | 30 | 93.3 | (79–98.2) | 41 | 85 | (72–93.1) | 0.453 |
|  | >30 | 25 | 20 | (8.9–39) | 53 | 17 | (9.2–29) | 0.759 |

CI: Confidence Interval; Ct: Cycle Threshold; n: number of SARS-CoV-2 positive samples tested; \*: significant at P<0.05 for difference in POCT sensitivity between pre-Omicron and Omicron variants using Fisher's exact test.

### Limit of Detection Analysis of POCTs With All Variants

Probit regression analyses was performed using data from all 149 SARS-CoV-2 clinical specimens to determine the 95% LoD of each test (Fig. 1). The molecular POCTs, the ID NOW and Check IT, had 95% LoDs of 34.7 and 39.3 Ct values, respectively, highlighting their ability to detect low quantities of virus. RADTs had 95% LoDs ranging between 25.0 to 28.4 (Table A1).

**FIG 1.**
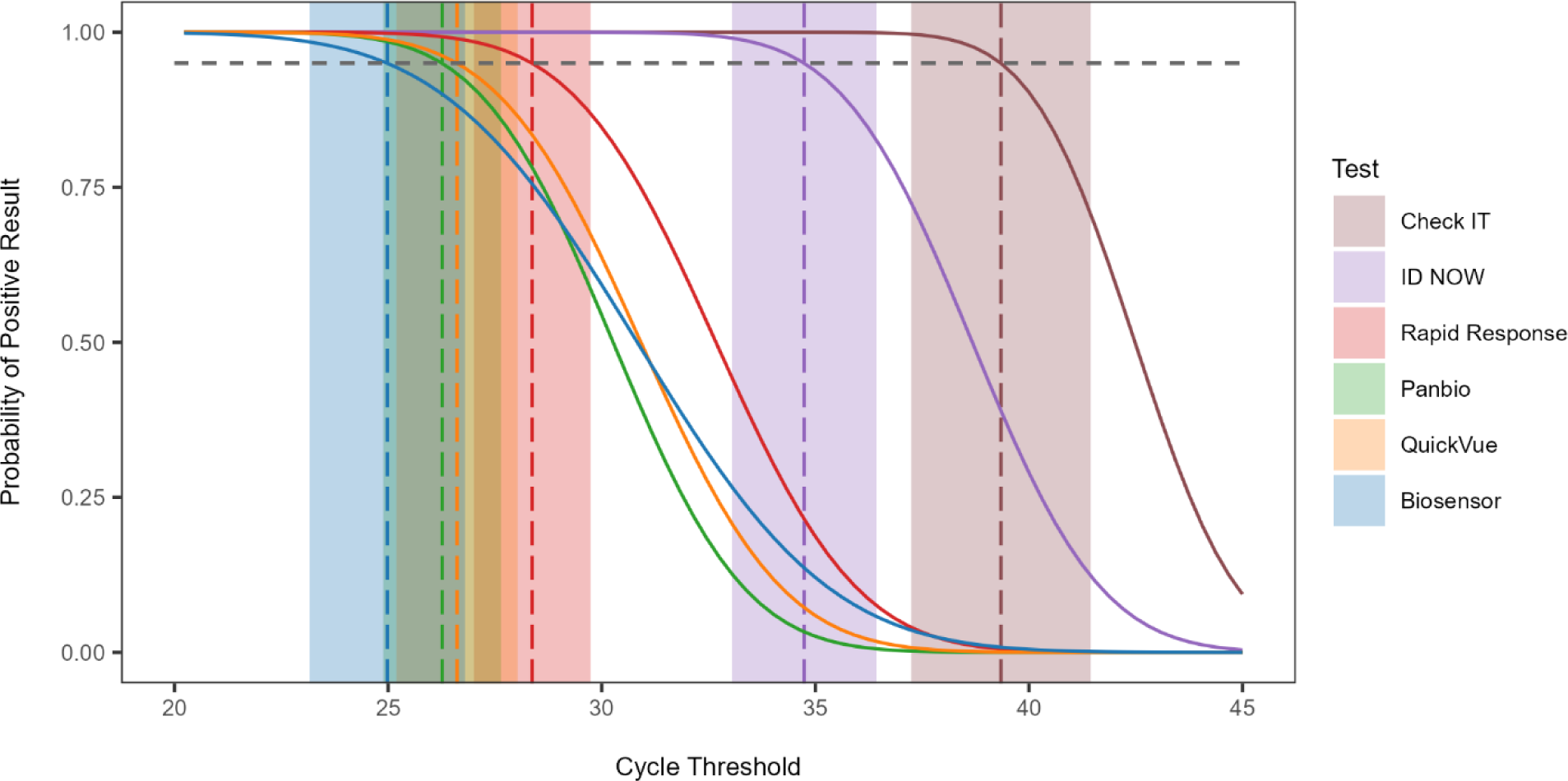
Probit regression analysis showing the 95% limit of detection of six SARS-CoV-2 point-of-care tests based on testing outcomes of 149 positive SARS-CoV-2 clinical samples. The cycle threshold of the sample, determined using the RT-qPCR reference standard test result, is plotted against a positive (1) or negative (0) test outcome. The horizontal dashed line indicates the 95% probability of a positive test result while the vertical dashed lines show the 95% limit of detection of each test with shaded areas representing their 95% confidence interval.

### Limit of Detection Analysis of POCTs For Pre-Omicron and Omicron Variants

A probit regression model was used to assess if the limit of detection of each test differed when stratified by variant (pre-Omicron vs Omicron). The 95% LoDs of all the POCTs were similar, regardless of the variant, as demonstrated by the sizeable overlap between their confidence intervals (Fig. 2). Due to a poor model fit from lack of false negative results, it was not possible to determine the 95% LoD of the Check IT for the pre-Omicron and Omicron variant samples, suggesting that performance of this molecular test is on par with the reference test.

**FIG 2.**
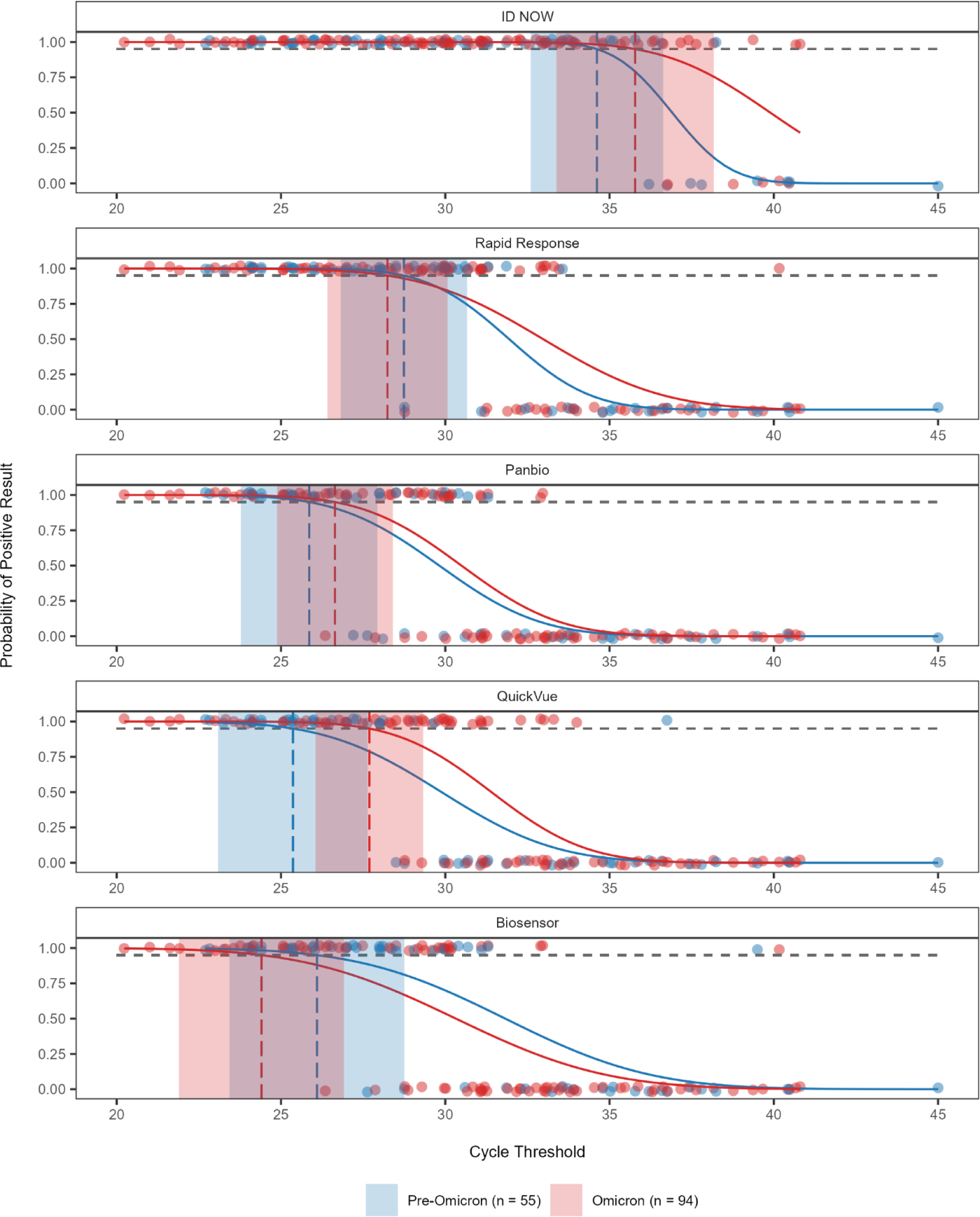
Probit regression analysis with the 95% limit of detection of five SARS-CoV-2 point-of-care tests grouped by variant, based on testing outcomes of 149 positive SARS-CoV-2 clinical samples. Results for pre-Omicron and Omicron samples are represented in blue and red, respectively. The cycle threshold of the sample, determined by the RT-qPCR reference standard test result, is plotted against a positive (1) or negative (0) outcome of the test. The horizontal dashed line indicates the 95% probability of a positive test result while the vertical dashed lines show the 95% limit of detection of each test with shaded areas representing their 95% confidence interval.

## DISCUSSION

POCTs have been central to the COVID-19 pandemic response, particularly following the appearance of the Omicron variant in late 2021 (2, 3). As new variants emerge, it is important to monitor the performance of POCTs to ensure that test results continue to be reliable. Consequently, the purpose of this study was to evaluate six widely-distributed Health Canada-approved POCTs using COVID-19 clinical samples incorporating pre-Omicron and Omicron variants (Alpha to XBB.1.5) to determine if test characteristics differed over time. Overall, our assessment revealed consistent and reliable performance across the variants tested, affirming manufacturer’s claims that there is no notable decrease in clinical efficacy. The molecular tests, ID NOW and Check IT, demonstrated the highest sensitivity, and can provide RT-qPCR level quality in settings where centralized testing is unavailable (30).

To our knowledge, only two studies, conducted by Stokes et al. (31, 32), have assessed the performance characteristics of the ID NOW in detecting both pre-Omicron and Omicron variants. Per Stokes et al. (31, 32), for symptomatic patients, the ID NOW demonstrated improved sensitivity against Omicron (91.6–96.0%) compared to pre-Omicron (90.0–92.5%). This aligns with the significant increase in test sensitivity against high Ct clinical Omicron samples noted in this study. Potential explanations for this finding may be attributed to various factors including changes in testing procedures over time, patient infection stage, or sample storage and quality. Studies have also reported decreased ID NOW performance with higher Ct samples (33–35). In contrast to traditional RT-qPCR, the ID NOW employs LAMP-based technology that targets a large RNA fragment (>1 kb). Therefore, the device may exhibit reduced sensitivity with fragmented samples from long-term storage, which may be a confounding factor in clinical LAMP validation studies. Additionally, the ID NOW targets RNA-dependant RNA polymerase, an uncommon target among COVID-19 POCTs and RT-qPCR assays. This feature may cause the ID NOW to be affected uniquely by variant mutations as compared to other tests, and the Ct value from the reference assay (targeting N2) may not accurately reflect the abundance of target RNA in each sample.

The Check IT has only been independently evaluated in one other publication to date. In a study by Zahavi et al. (8), the device exhibited a sensitivity of 91.1% using samples collected before the emergence of Omicron. This is consistent with the high sensitivity of the Check IT when using both pre-Omicron and Omicron variants observed in this study, with the LoD comparable to that of laboratory-developed qRT-PCR tests.

Using the WHO guidelines for RADT evaluation and clinical relevance (27), at high viral loads (Ct ≤ 30) all RADTs met the criteria for acceptable clinical sensitivity (> 80%). In agreement with previous reports and manufacturer claims, test sensitivity of RADTs decreased markedly at low viral loads (Ct > 30) regardless of variant. It is important to note that comparing LoDs and sensitivity across studies, especially for RADTs, is often challenging due to differences in experimental design. For example, reference Ct values can differ greatly based on methodology and gene target. Recently, media reported on possible issues in the design of the BTNX validation studies used for regulatory approval (36); however, this study corroborates manufacturer claims and has identified no appreciable differences in analytical test performance between the BTNX and the other RADTs investigated in this study.

A meta-analysis by Mohammadie et al. (37) on RADT performance revealed a pooled sensitivity of 67% and a non-significant reduction in overall RADT sensitivity against Omicron. While these general trends provide valuable insights into overall RADT performance, the diversity of targets needs to be considered. Although as a group RADTs demonstrate compatibility with emerging variants, each test recognizes unique viral epitopes and should also be investigated individually.

The QuickVue RADT showed significantly improved sensitivity against low viral load Omicron samples, consistent with findings by Sugiharto et al. (24); however, work by Rao et al. (38) showed no change in QuickVue sensitivity against Omicron variants. All other RADTs evaluated in this study showed no significant change in test performance against Omicron variants. Previous studies evaluating RADTs have consistently reported similar or reduced clinical sensitivity with Omicron as compared to early pandemic variants (21, 37, 39). For example, multiple studies investigating the Abbott Panbio using cultured virus (40) and clinical samples (23, 41) have shown no sensitivity loss against Omicron. Alternatively, work by Bekliz al. (19) reported a significant reduction in Panbio sensitivity with Omicron BA.1 compared to Delta. Biosensor demonstrated comparable performance with cultured Omicron variants (40, 42), but in other studies exhibited reduced clinical sensitivity (19, 20). To our knowledge, this is the first study investigating the sensitivity of the BTNX Rapid Response against multiple SARS-CoV-2 variants including Omicron.

A limitation of this study is the use of anonymized remnant clinical samples instead of clinical patient swabs. The use of clinical swabs was not feasible as multiple variants investigated in this study are no longer circulating and this study required a large number of paired samples to evaluate multiple technologies. Due to the anonymization of samples used in this study, we were not able to incorporate information that may affect test sensitivity or specificity, such as vaccination status or patient clinical presentation, into our analyses (13, 14, 39).

Unlike many studies that primarily focused on comparing BA.1 exclusively with the Delta variant, our research included a representative range of both pre-Omicron and Omicron subvariants (Alpha to XBB.1.5). Considering both the overall sensitivity and limit of detection, we observed no decline in test performance against Omicron variants; however, it is important to continue to evaluate these parameters as new variants emerge. Sustained post-market surveillance efforts are required to proactively identify issues that could impact decentralized testing for COVID-19.

## ETHICS APPROVAL

No ethics were required under Article 2.4 and Article 2.5 of the *Tri-Council Policy Statement Ethical Conduct for Research Involving Humans* (26) since remnant clinical specimens were used for the purpose of quality assurance.

## SUPPLEMENTAL MATERIAL

**Table A1.** Probit regression calculated 95% Limit of Detection of six COVID-19 point-of-care tests based on 149 SARS-CoV-2 positive clinical samples.

| Test | Predicted 95% LoD (Ct) 95% CI<sup>1</sup> |  |  |  |  |  |
| --- | --- | --- | --- | --- | --- | --- |
|  | All (n=149) |  | Pre-omicron (n=54) |  | Omicron (n=95) |  |
| Molecular Tests |  |  |  |  |  |  |
| ID NOW | 34.7 | (33.1, 36.4) | 34.6 | (32.6, 36.6) | 35.8 | (33.4, 38.2) |
| Check IT | 39.3 | (37.2, 41.4) | N/A<sup>2</sup> |  | 38.7 | (35.6, 41.8) |
| Rapid Antigen Tests |  |  |  |  |  |  |
| Panbio | 26.3 | (24.9, 27.6) | 25.9 | (23.8, 27.9) | 26.6 | (24.9, 28.4) |
| Rapid Response | 28.4 | (27.0, 29.7) | 28.7 | (26.8, 30.7) | 28.2 | (26.4, 30.1) |
| QuickVue | 26.6 | (25.2, 28.0) | 25.4 | (23.1, 27.6) | 27.7 | (26.1, 29.3) |
| Biosensor | 25.0 | (23.2, 26.8) | 26.1 | (23.4, 28.8) | 24.4 | (21.9, 26.9) |

<sup>1</sup>95% Wald confidence intervals; <sup>2</sup>N/A since insufficient data, n - number of SARS-CoV-2 positive samples.

## Data Availability

All primary research data used to generate this manuscript are included in-text and within Supplementary Data 1.

